# Vaccination boosts protective responses and counters SARS-CoV-2-induced pathogenic memory B cells

**DOI:** 10.1101/2021.04.11.21255153

**Authors:** Pankaj Kumar Mishra, Natalie Bruiners, Rahul Ukey, Pratik Datta, Alberta Onyuka, Deborah Handler, Sabiha Hussain, William Honnen, Sukhwinder Singh, Valentina Guerrini, Yue Yin, Hannah Dewald, Alok Choudhary, Daniel B. Horton, Emily S. Barrett, Jason Roy, Stanley H. Weiss, Patricia Fitzgerald-Bocarsly, Martin J. Blaser, Jeffrey L. Carson, Reynold A. Panettieri, Alfred Lardizabal, Theresa Li-Yun Chang, Abraham Pinter, Maria Laura Gennaro

## Abstract

Much is to be learned about the interface between immune responses to SARS-CoV-2 infection and vaccination. We monitored immune responses specific to SARS-CoV-2 Spike Receptor-Binding-Domain (RBD) in convalescent individuals for eight months after infection diagnosis and following vaccination. Over time, neutralizing antibody responses, which are predominantly RBD specific, generally decreased, while RBD-specific memory B cells persisted. RBD-specific antibody and B cell responses to vaccination were more vigorous than those elicited by infection in the same subjects or by vaccination in infection-naïve comparators. Notably, the frequencies of double negative B memory cells, which are dysfunctional and potentially pathogenic, increased in the convalescent subjects over time. Unexpectedly, this effect was reversed by vaccination. Our work identifies a novel aspect of immune dysfunction in mild/moderate COVID-19, supports the practice of offering SARS-CoV-2 vaccination regardless of infection history, and provides a potential mechanistic explanation for the vaccination-induced reduction of “Long-COVID” symptoms.

---

Since December 2019, the novel severe acute respiratory syndrome coronavirus 2 (SARS-CoV-2) has infected >130 million people and caused about three million deaths worldwide (https://coronavirus.jhu.edu/map.html). The antibody response to SARS-CoV-2 infection has been a major research focus (examples are ^1-3^), as it is directly relevant to diagnosis, seroprevalence studies, vaccine development, and immunotherapy ^4,5^. One year into the pandemic, our knowledge of antibody longevity and function and antibody-producing cell lineages in this infection is still evolving. Moreover, the ongoing SARS-CoV-2 vaccination campaign worldwide, including two mRNA vaccines [Moderna (mRNA-1273) or Pfizer–BioNTech (BNT162b2)] ^6,7^, raises questions about the properties and duration of humoral and cellular immune responses to vaccination. While studies are being rapidly published [examples are ^8-10^], much remains to be learned, particularly in terms of determining how prior SARS-CoV-2 infection impacts the response to the vaccines and comparing responses to vaccination versus infection.

Here we examine SARS-CoV-2-specific antibody and memory B cell responses of 83 Rutgers University employees who were infected with SARS-CoV-2 during the first wave of the COVID-19 pandemic in New Jersey (March-June 2020). We also characterize antibody and memory B cell responses in a subset of this cohort that received full (two-dose) administration of SARS-CoV-2 mRNA vaccines. We compare the vaccine responses in these subjects to the corresponding pre-vaccination responses to infection and to the vaccine responses in a comparator group of infection-naïve subjects.

SARS-CoV-2 infection had been confirmed in 81 of 83 study subjects by virus-specific quantitative RT-PCR [two subjects were diagnosed by their physicians based on household exposure history, symptoms, and chest x-ray findings]. Infection induced mild to moderate symptoms in the vast majority of subjects, with only 5 (6%) reporting COVID-19-related hospitalization (for demographics and clinical information, see **Table S1** and **Fig. S1A**). We first analyzed the plasma samples of all 83 infected subjects for levels of antibodies directed to the receptor binding domain (RBD) of the S1 subunit of the SARS-CoV-2 Spike protein. The coronavirus Spike protein mediates viral entry into host cells through an interaction of RBD with the ACE-2 host receptor ^11^. We chose RBD as target antigen of the antibody response because it is immunodominant and features limited sequence conservation among coronaviruses ^12^, which minimizes the potential detection of cross-reactive antibodies. We found that detection of RBD-specific IgG antibodies clearly separated subjects who had tested positive to SARS-CoV-2 PCR (n = 83) from negative control subjects [pre-COVID-19 (n = 104) and SARS-CoV-2 PCR-negative subjects (n = 103) that remained SARS-CoV-2 PCR-negative for at least 16 weeks after the blood draw tested in the figure] (**Fig. 1A**). To monitor antibody responses over time, we retained 22 subjects for serial blood draws (monthly for 3 months and then bimonthly) over an eight-month period (April-December 2020). The demographics and clinical severity of these 22 participants did not substantively differ from those of the 83 subjects, except that they included no participants who had been hospitalized for COVID-19 (**Table S2**). The trajectories of RBD-specific IgG, IgM, and IgA antibodies were heterogeneous (**Fig. 1B, Fig. S1B-D**). In particular, the IgG response declined over time in 16 subjects (73%) and remained stable or increased in 6 subjects (27%) (**Fig. 1B**). We also analyzed the virus neutralization activity of the plasma collected at the first and last study visit, using an assay with replication-competent SARS-CoV-2 virus (**Fig. S1E)**. Neutralization activity decreased in most subjects (n = 15, 68%) (**Fig. 1C**) and positively correlated with RBD-specific antibody titers (r = 0.71; p<0.0001) (**Fig. S1F**), as previously reported [for example, ^13^]. To directly test the link between RBD specificity and the antibody-mediated ability of plasma to neutralize the virus, we depleted RBD-specific antibodies from seropositive plasma samples, and, as a comparator, we also depleted viral Nucleocapsid (N)-specific antibodies, and then tested the effects on neutralizing activity. We first confirmed by ELISA that adsorption with either antigen resulted in depletion of the corresponding specific IgG, with little (if any) reduction in antibody binding to the second (non-absorbing) antigen (**Fig. 1D-E**). The neutralizing activity of the plasma was abrogated only when the samples were preabsorbed with RBD, but not when they were preabsorbed with N (**Fig. 1F**). This provides direct evidence that most (if not all) of the plasma neutralizing activity resides in the RBD-specific antibodies. Collectively, the data show that, at least in subjects that have experienced mild symptoms, circulating levels of RBD-specific IgG tend to decrease over time, with concurrent reduction in plasma neutralizing titers. Decreases in circulating antibodies below levels of detection could eventually lead to an underestimation of the prevalence of SARS-CoV-2 infection using serological methods. In addition, our antibody depletion approach, which has been utilized in COVID-19 research very rarely [we find it used only in two reports ^10,14^], shows that the neutralizing activity of convalescent plasma mostly resides in the RBD-specific antibodies. This result provides a framework for monoclonal antibody studies directed toward antibody-based therapeutics [examples are ^15,16^] and for the rigorous assessment of the value of COVID-19 plasma therapy, which remains controversial ^17^, in part due to the poor characterization of the products used.

**Fig. 1.**
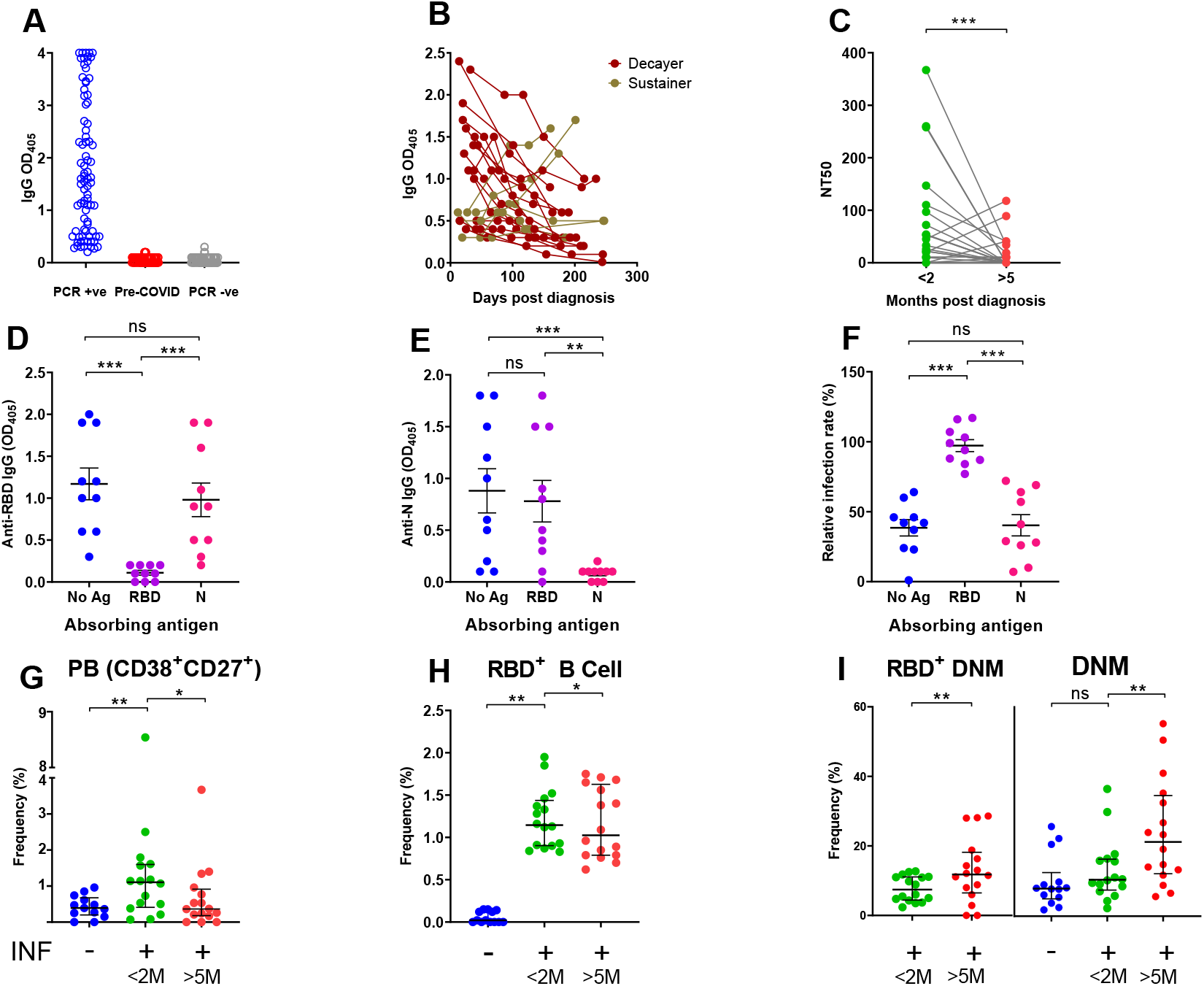
Temporal trajectories of antibody levels, plasma neutralization activity, and memory B cell responses to SARS-COV-2 infection. (**A**,**B**) Receptor Binding Domain (RBD)-specific antibody binding. (**A**) Anti-RBD IgG antibody levels in SARS-CoV-2 PCR-positive subjects (n = 83, blue circles); pre-COVID-19 samples (n = 104; red circles); subjects SARS-CoV-2 PCR-negative for at least 16 weeks after blood draw (n = 103 subjects; grey circles). Each circle indicates one subject. (**B**) Temporal trajectory of IgG antibody levels in SARS-CoV-2-infected subjects. Symbol colors distinguish decayers (n=16) showing downward trajectories (dark red) and sustainers (n=6) showing stable or upward trajectories (dark yellow). (**C**) Neutralizing titers expressed as NT50 (reciprocal dilution of plasma yielding 50% virus neutralization), as obtained with samples collected <2 months (green circles) or >5 months (red circles) post-SARS-CoV-2 diagnosis from the same 22 subjects as in **B**. Each pair of circles connected by a line represents one subject. (**D-F**) Selected plasma samples (n = 10) exhibiting NT50 >80 were depleted of either RBD or N-specific antibodies by pre-incubating them with either recombinant RBD or N (Nucleocapsid) proteins and then performing ELISA for IgG binding to RBD (panel **D**) or N (panel **E**) as solid-phase antigen. (**F**) Neutralization activity of pre-absorbed plasma samples, as in **D-E**. Relative infection rates were calculated as in Methods and Fig. S1. **(G-I)** Frequency (%) of selected B cell compartments: plasmablasts (PB) (panel **G**), RBD-specific (RBD^+^) B cells (panel **H**), and double negative memory B cells (DNM) (panel **I**). <2M and >5M, as in panel **C**. For the gating strategy, see supplementary **Fig. S2A-H**. The entire dataset is found in supplementary Table 3. In all panels, the individual circle represents one subject, and the solid black lines represent median with interquartile range. Statistical analysis was performed either with Mann-Whitney *U-*test for unpaired data or Wilcoxon matched-pairs signed rank test for paired samples (**, p ≤ 0.01; ***, p ≤ 0.001; ns, non-significant, p > 0.05).

Since the progressive decrease of protective antibodies in the circulation raises the important question of whether immune protection against SARS-CoV-2 infection also wanes over time, we next analyzed the memory B cell response in the same subjects. To accomplish this, we developed a multicolor flow cytometry panel to measure frequencies of circulating B cells (CD19^+^CD20^+^) and B cell subsets including plasmablasts (CD27^+^CD38^hi^), naïve (CD27^-^IgD^+^) and memory B cell compartments [non-switched memory (CD27^+^IgD^+^), switched memory (CD27^+^IgD^-^), and double negative memory (CD27^-^IgD^-^)] (**Fig. S2A-H**). We also evaluated frequencies of RBD-specific B cells (RBD-tetramer-positive CD19^+^CD20^+^) (**Fig. S2D**), which we further subdivided into memory compartments (**Fig. S2F)**. The results of all analyses are shown in **Tables S3-S4**. Comparing results obtained from our previously infected subjects with a group of 13 SARS-CoV-2 infection-naive individuals [pre-COVID (n = 5) and SARS-CoV-2 PCR-negative samples (n = 8)] showed that SARS-CoV-2 infection correlated with a transient increase of plasmablast frequencies (**Fig. 1G**) and the appearance of antigen-specific (RBD^+^) B cells (**Fig. 1H**). Plasmablast frequencies are known to increase in response to acute infections, including SARS-CoV-2 [for example, ^18,19^]. Among circulating non-plasmablasts, changes of frequencies in circulating naïve and switched memory B cells reflect B cell remodeling in response to recent infection (**Table S3**). Analysis of RBD^+^ memory B cells in the first-vs last-visit samples showed stable frequencies over time, indicating a durable memory response (**Fig. 1H**). Notably, the frequency of an RBD^+^ double negative memory (DNM) B cell subset (CD27^-^IgD^-^) increased between the two study visits (**Fig. 1I**, left panel). An increase was also observed with the total pool of DNM B cells (**Fig. 1I**, right panel). The DNM B cell subset, which increases with age ^20^, is considered a component of the B cell memory compartment despite the absence of the CD27 memory marker, since it carries signatures of antigen-experienced B cells in terms of surface phenotype, proliferation response, and patterns of somatic hypermutations ^21^. DNM B cells likely constitute a heterogeneous B cell subset, as they have been described as exhausted / prematurely senescent B cells in HIV infection and other diseases characterized by chronic immune activation ^22,23^, or as producing autoantibodies in autoimmune diseases such as systemic lupus erythematosus ^24^. Interestingly, SARS-CoV-2-induced exhaustion and senescence phenotypes have been previously reported for T cells ^25^. Thus, our data imply that immune exhaustion, which presumably correlates with the degree of inflammation and disease severity, occurs in both B cells and T cells during SARS-CoV-2 infection. Moreover, increased frequencies of DNM B cells may correlate with the production of autoantibodies reported in pediatric and adult COVID-19 patients ^26-28^. Further studies of DNM B cell phenotypes, function, and fate across the COVID-19 severity spectrum are warranted.

Some subjects in our longitudinal cohort received COVID-19 vaccination (mRNA vaccines) in December 2020 - February 2021, providing the opportunity to determine whether re-exposure to antigen via vaccination induces recall humoral and B cell responses. We measured antibody and B cell responses to full vaccination (two doses of COVID-19 mRNA vaccines) in 12 cohort subjects. As infection-naïve comparators, we also tested 8 subjects who were SARS-CoV-2 PCR-negative and seronegative prior to vaccination. We found that, in both groups, vaccination induced very vigorous antibody responses, which were much higher in the previously infected than infection-naïve individuals (**Fig. 2A**). Moreover, IgG titers were much higher in infection-naïve vaccinated than in infected unvaccinated subjects at the first study visit (i.e., when they were recently infected) (**Fig. 2B**). Similar to the effect on antibody responses, vaccination induced stronger neutralizing activity in the previously infected group (**Fig. 2C**). Moreover, vaccination of the infection-naïve group led to a stronger neutralizing activity than did recent infection alone (**Fig. 2D**). We also found that vaccination of the infected group led to higher RBD-specific memory B cell frequencies than vaccination of the infection-naïve group (**Fig. 2E**). Collectively, these results lead to three clear conclusions. First, administration of COVID-19 RNA vaccines induces vigorous humoral and B cell responses, regardless of infection history. Second, vaccination elicits a strong recall response in subjects previously exposed to the virus. Third, vaccination elicits stronger responses than SARS-CoV-2 infection, at least when it presents with mild or moderate symptoms. Analysis of the memory B cell pool showed that, in all subjects, vaccination had no effect on circulating plasmablast frequencies and was associated with remodeling and/or redistribution in the periphery of B cell memory compartments (increased naïve and decreased switched memory B cells) (**Table S4**). Notably, following vaccination, the frequencies of DNM B cells decreased in previously infected individuals (but not in infection-naïve subjects); although sample sizes are small, the differences between paired samples were statistically significant (**Fig. 2F**). These results indicate that the vaccine response counters the infection-induced increased production of dysfunctional and potentially pathogenic immune cells (**Fig. 1I**).

**Fig. 2.**
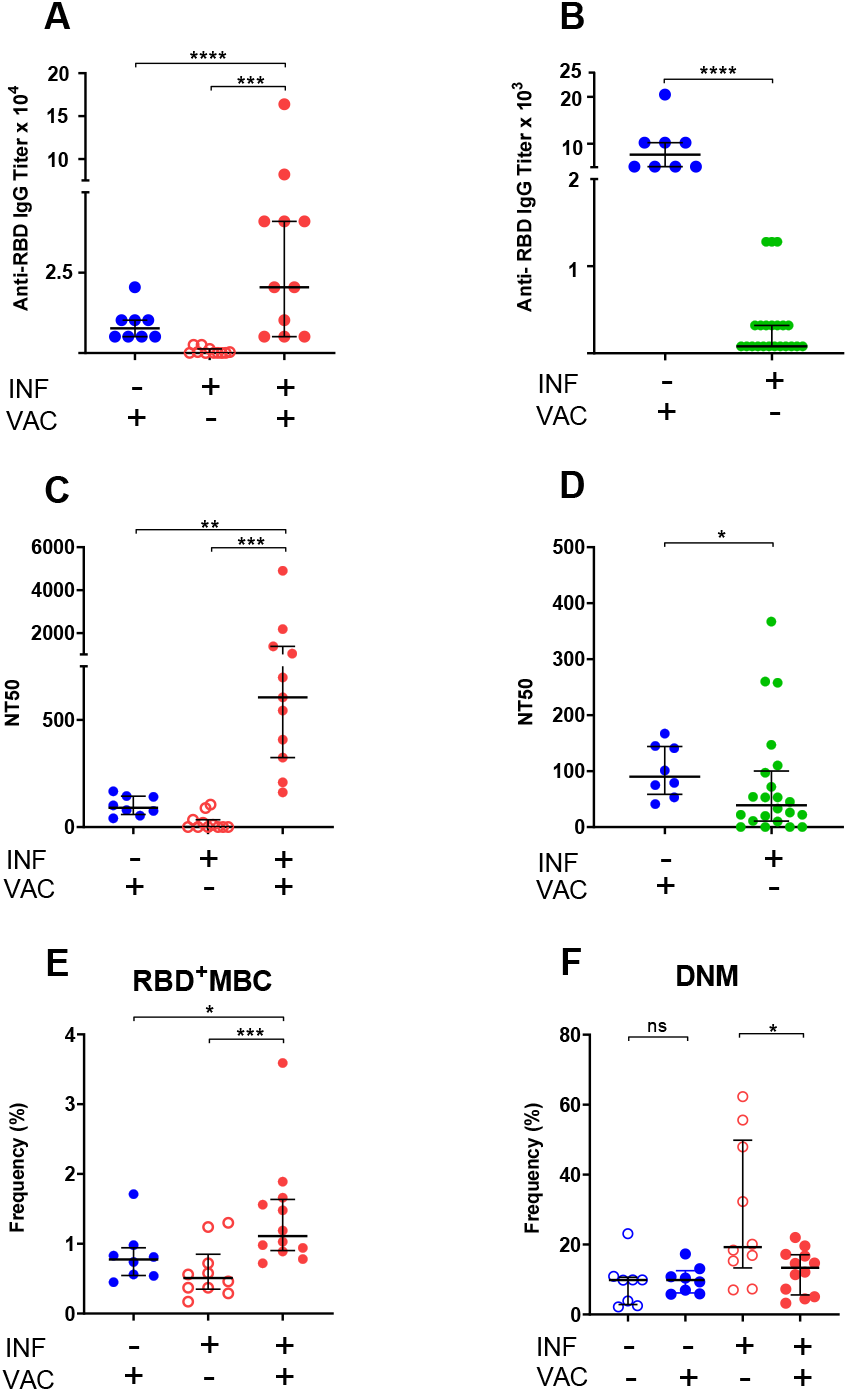
Humoral and B cell responses to COVID-19 mRNA vaccination. Responses to COVID-19 RNA vaccines in previously infected study subjects and infection-naïve comparators were measured. Receptor Binding Domain (RBD)-specific IgG antibody binding, plasma neutralizing activity, and B cell frequencies were assessed as in **Fig. 1**. Infection (INF) and vaccination (VAC) status are indicated by the corresponding (+) and (-) signs. (**A**,**B**) Titers of RBD-specific IgG by ELISA. (**C**,**D**) NT50 determinations. Pre-vaccination samples of SARS-CoV-2-infected subjects were either those collected at the last visit before vaccination to determine effect of vaccination (panels **A**,**C**) or those collected at the first visit post-diagnosis to compare responses to recent infection and recent vaccination (panels **B**,**D**). (**E**) Frequency of RBD-specific memory B cells (RBD^+^ MBC). (**F**) Frequency of total double-negative memory B cells (DNM, CD27^-^ IgD^-^). The entire B cell dataset is found in supplementary Table 4. In all panels, data are shown as scatter plots; each circle represents one study subject; the solid black lines represent the median and interquartile range. Statistical analysis was performed either with Mann-Whitney *U-*test for unpaired data or Wilcoxon matched-pairs signed rank test for paired samples (*, *p* ≤ 0.05; **, *p* ≤ 0.01).

In conclusion, in subjects who have experienced mild SARS-CoV-2 infection, we demonstrate that the decrease of circulating protective antibodies over time is accompanied by durable memory responses that are competent to induce potent recall responses upon re-exposure to antigen. Our work confirms studies showing potent immune responses induced by anti-SARS-CoV-2 vaccines [for example, ^29^]. In particular, we show that the humoral and B cell responses to vaccination are more vigorous than those induced by recent infection alone (at least when symptoms are mild/moderate), supporting the current practice in the USA of offering SARS-CoV-2 vaccination regardless of infection history. The direct demonstration that the neutralizing activity of plasma resides predominantly in the RBD antibody specificities provides a molecular foundation to the prior correlations between antibody binding and antibody-mediated neutralizing activity and to the evaluation of antibody-based COVID-19 therapies. The accumulation of DNM B cells may explain the observed production of autoantibodies in COVID-19 patients and/or reveal the accumulation of exhausted/prematurely senescent B cells, which complements the known SARS-CoV-2-induced T cell exhaustion/senescence. Furthermore, while previous reports correlate DNM B cells with severe COVID-19 ^30,31^, we find that frequencies of DNM B cells increase over time in subjects who experienced relatively mild symptoms. Since accumulation of these potentially pathogenic B cell subset in lymphatic organs and in the periphery correlates with loss of germinal centers and follicular B cells in severe COVID-19 patients ^30^, our finding strongly suggests that immune dysfunction can also be associated to mild disease and can worsen even after symptom resolution. Unexpectedly, vaccination counters this aspect of immune dysregulation. The still anecdotal reports of symptomatic relief after vaccination in persons with post-acute sequelae of SARS-CoV-2 infection (for example, ^32^) may find mechanistic grounds in these findings.

## Materials and Methods

### Study participants

Eighty-three convalescent individuals with a history of SARS-CoV-2 infection were enrolled between April 1 and June 17, 2020 from among Rutgers University employees to monitor virus-specific immune responses. Participants showed proof of positive results of SARS-CoV-2 PCR assays that had received Emergency Use Authorization from the Food and Drug Administration with the exception of two subjects who received a clinical diagnosis only. All participants self-reported the date of symptom onset and consented to blood draws as well as the collection of demographics and clinical symptoms. Of 83 participants, 22 were retained for 3-6 study visits over a period of 7 months (once a month for the first 3 months, and then once every two months). As negative control for SARS-CoV-2 antibody testing, we used 104 stored serum/plasma samples collected prior to the COVID-19 pandemic (Institutional review board of the Rutgers New Jersey Medical School, Pro0119980237 and Pro20150001314) and 103 serum samples obtained during the pandemic from subjects that remained SARS-CoV-2 PCR-negative and seronegative for at least 16 weeks following the test blood draw ^33^ (ClinicalTrials.gov registration number NCT04336215). Among study participants who were vaccinated with either Moderna or Pfizer mRNA vaccines between mid-December 2020 and mid-February 2021, 12 donated blood at least two weeks after the second vaccine dose between February 22 and March 1, 2021. Between December 22, 2020 and February 11, 2021, blood pre-and post-vaccination was also obtained from 8 subjects who were SARS-CoV-2 PCR-negative and seronegative prior to vaccination.

### Antibody binding by enzyme-linked immunosorbent assay (ELISA)

96-well ELISA plates (Nunc MaxiSorp, ThermoFisher, Rochester NY) were coated with 2 µg/ml recombinant SARS-CoV-2 RBD overnight at 4°C. Plates were washed with washing buffer (1 X PBS containing 0.05% Tween 20) (Sigma-Aldrich, St. Louis MO) and incubated with 2% BLOTTO (Nestle Carnation, US) in PBS for 30 min at 37°C. Plasma was heat-inactivated at 56°C for 1 hour prior to use. After blocking, diluted plasma was added in blocking buffer and incubated for 1 hour at 37°C. Antigen-specific IgG was detected by adding alkaline phosphatase-conjugated goat anti-human IgG (Jackson ImmunoResearch, West Grove PA). ELISA plates were developed using phosphate substrate (Sigma-Aldrich) and the reaction was stopped with 1M NaOH. The ELISA protocol was performed using a BioTek EL406 combination washer dispenser, and absorbance (OD_405_nm) was measured using a BioTek Synergy Neo2 microplate reader (BioTek, Winooski VT). Each ELISA plate contained positive and negative serum/plasma controls and background control wells without primary antibody. Each sample was tested in duplicate. End-point titers were plotted for each sample using background-subtracted data. All work involving blood products from SARS-CoV-2-infected subjects were performed in a biosafety level 2+ (BSL-2+) laboratory utilizing protocols approved by the Rutgers Institutional Biosafety Committee.

### Expression and purification of Recombinant SARS-COV-2 S1 RBD Protein

A DNA fragment encoding RBD (Spike residues aa. 319 to aa. 537) was amplified and cloned into the eukaryotic expression vector pcDNA3.1 (Addgene, Watertown MA). Purified plasmid was transfected into 293F cells using the Expi293 Expression system (Thermo Fisher Scientific, Waltham MA), according to the manufacturer’s protocol. Supernatants were collected on day 3 post-transfection, purified by HisPur™ Cobalt Resin (Thermo Fisher Scientific) and eluted with 200mM imidazole. Purified protein was subsequently dialyzed against PBS at 4°C. Absorbance (OD_280_nm) was determined by Nanodrop reading and concentrations were calculated using ExPASy Proteomics calculator. Molecular weights were adjusted to account for the number of N-linked glycosylation sites to determine the final concentration.

### Absorption of convalescent plasma with SARS-CoV-2 antigens

ELISA 96-well microtiter plates were coated with 500 ng/well of SARS-CoV-2 RBD or N proteins at 4°C overnight. Coated plates were washed three times with washing buffer and blocked with PBS containing 1% BSA (Sigma-Aldrich) for 30 min at 37°C. After washing, plasma samples were diluted 1:10 in PBS containing 1% BSA (Sigma-Aldrich) and incubated up to overnight at 4°C for each cycle. Absorption was repeated at least four times utilizing fresh antigen-coated plates at each cycle. To monitor depletion of antigen-specific antibodies, RBD-and N-specific IgG titers of untreated and absorbed samples were determined as described above, prior to use in neutralization assays.

### Cell lines

Vero E6 were obtained from the American Type Culture Collection (ATCC; Manassas VA) and HeLa cells stably expressing ACE2 (HeLa-ACE2) were obtained from Dennis Burton at the Scripps Research Institute ^34^. All cell lines were maintained in high-glucose Dulbecco’s modified Eagle’s medium (DMEM; Corning, Manassas VA) supplemented with 10% fetal bovine serum (FBS; Seradigm, Radnor PA), 2mM L-glutamine (Corning),1% penicillin/streptomycin (Corning), and incubated in humidified atmospheric air containing 5% CO_2_ at 37°C.

### SARS-CoV-2 virus

The virus stock of mNeonGreen (mNG) SARS-CoV-2 was obtained from Pei-Yong Shi at the University of Texas Medical Branch at Galveston. The virus stock was produced using the virus isolate of the first patient diagnosed in the US, in which the ORF7 of the viral genome was replaced with the reported mNG gene ^35^. Propagation of viral stocks was performed with Vero E6 cells using 2% FBS. The virus titers were determined by standard plaque assay utilizing Vero E6 cells ^36^ and recorded as plaque forming units per milliliter (PFU/mL). All work involving live SARS-CoV-2 were performed in a biosafety level 3 (BSL-3) laboratory utilizing protocols approved by the Rutgers Institutional Biosafety Committee.

### SARS-CoV-2 neutralization assay

HeLa-Ace2 cells were seeded in 96-well black optical-bottom plates at a density of 1 × 10^4^ cells/well in FluoroBrite DMEM (Thermo Fisher Scientific) containing 4% FBS (Seradigm), 2mM L-glutamine (Corning),1% penicillin/streptomycin (Corning) and incubated overnight at 37°C with 5% CO_2_. On the following day, each sample was subjected to two-fold serial dilution in DMEM without FBS, and incubated with mNG SARS-CoV-2 at 37°C for 1.5 h. The virus-plasma mixture was transferred to 96-well plates containing Hela-Ace2 cells at a final multiplicity of infection (MOI) of 0.25 (viral PFU:cell). For each sample, the starting dilution was 1:20 and the final dilution of 1:10,240. After incubating infected cells at 37°C for 20 h, mNG SARS-CoV-2 fluorescence was measured using a Cytation™ 5 reader (BioTek). Relative fluorescent units were converted to percent neutralization by normalizing the sample-treatment to non-sample-treatment controls and plotted with a nonlinear regression curve fit to determine the titer neutralizing 50% of SARS-CoV-2 fluorescence (NT_50_). Each patient sample was tested in duplicate. All plasma samples were heat-inactivated at 56°C for 60 min before testing.

### PBMC isolation and storage

Peripheral blood mononuclear cells (PBMC) were isolated by Ficoll density gradient centrifugation (Ficoll-Paque, GE Healthcare, Uppsala, Sweden), as described ^37^. Briefly, whole blood was diluted with equal volume of Roswell Park Memorial Institute Medium (RPMI; Corning) and layered on Ficoll-Paque (GE healthcare, USA). The gradient was centrifuged at 500 *× g* for 30 min at room temperature. Plasma was carefully removed, aliquoted, and stored at −80°C. The PBMC interface was collected, washed once, and counted using a hemocytometer. PBMCs were cryopreserved in liquid nitrogen in FBS containing 10% dimethyl sulfoxide (DMSO; Thermo Fischer Scientific) and stored until use.

### B cell immunophenotyping

Biotinylated RBD (Biolegend, San Diego CA) was mixed with streptavidin PerCP-Cy5.5. (Thermo Fischer Scientific) at 4:1 molar ratio for 1 hour at 4°C to form the RBD tetramer (RBD_4_-PercpCy5.5). PBMCs were incubated with the fixable viability stain 780 (BD Biosciences, Franklin Lakes NJ), followed by 10 min incubation with human Fc receptor blocking reagent (BD Biosciences). For the detection of antigen-specific B cells, PBMCs were incubated with the RBD tetramer and antibodies to CD19-BV700 (HIB19, BioLegend), CD20-PE-CF594 (2H7, BD Biosciences), CD27-PE (L128, BD Biosciences), CD38-BB515 (HIT2, BD Biosciences), IgM-BV605 (MHM-88, BioLegend), IgG-PE-Cy7 (G18-145, BD Biosciences). Additionally, cells were stained with APC-labelled antibodies against CD3 (UCHT1, Thermo Fisher Scientific), CD4 (OKT4, Thermo Fisher Scientific), CD14 (C1D3, Thermo Fisher Scientific), and CD16 (CB16, Thermo Fisher Scientific) to eliminate non-B cells. Antibody binding was allowed by incubating samples for 30 minutes on ice in the dark. Cells were washed twice with FACS buffer (0.1% BSA in PBS), fixed with 4% paraformaldehyde (PFA) for 20 min, and stored at 4°C overnight. For each sample, at least 250,000 events were acquired on a BD Fortessa X-20 flow cytometer (BD Biosciences).

### Statistical Analysis

All flow cytometry data were analyzed with FlowJo v12 software (FlowJo LLC, Ashland OR). Statistical analysis was performed with GraphPad Prism 8.4 (Graph Pad Software Inc., La Jolla CA). Data values are presented as median with interquartile range (IQR). Correlation analysis was performed using the non-parametric Spearman’s rank correlation. Statistical analysis was performed utilizing either Mann-Whitney U test for unpaired samples or Wilcoxon matched pairs signed rank test for paired samples. With all tests, p < 0.05 was considered significant.

## Data Availability

The data that support the findings of this study are all listed in the article and available from the corresponding author upon reasonable request.

## Acknowledgements

We thank the PHRI biosafety officers and RBHS Institutional Biosafety committee for fast-track review and approval of laboratory protocols and practices related to handling of SARS-CoV-2 and infected biospecimens; Daniel Fine and Steven Libutti for supporting the start of our COVID-19 work; Nancy Reilly and the entire Rutgers Corona Cohort team that established a cohort from which infection-naïve samples were obtained during the pandemic; Tracy Andrews of RUBIES for advice on statistical methods; David Alland for providing fluorescence-tagged SARS-CoV-2 virus; Dennis Burton and Pei-Yong Shi for providing biological reagents; and Helen Pickersgill at Life Science Editors for professional editorial services. This work was funded by NIH grants R01 HL149450, R01 HL149450-S1, U01 AI122285-S1, P30 ES005022, and UL1 TR003017.

