## Supplementary Figures and Tables for "Vaccination boosts protective responses and counters SARS-CoV-2-induced pathogenic memory B cells"

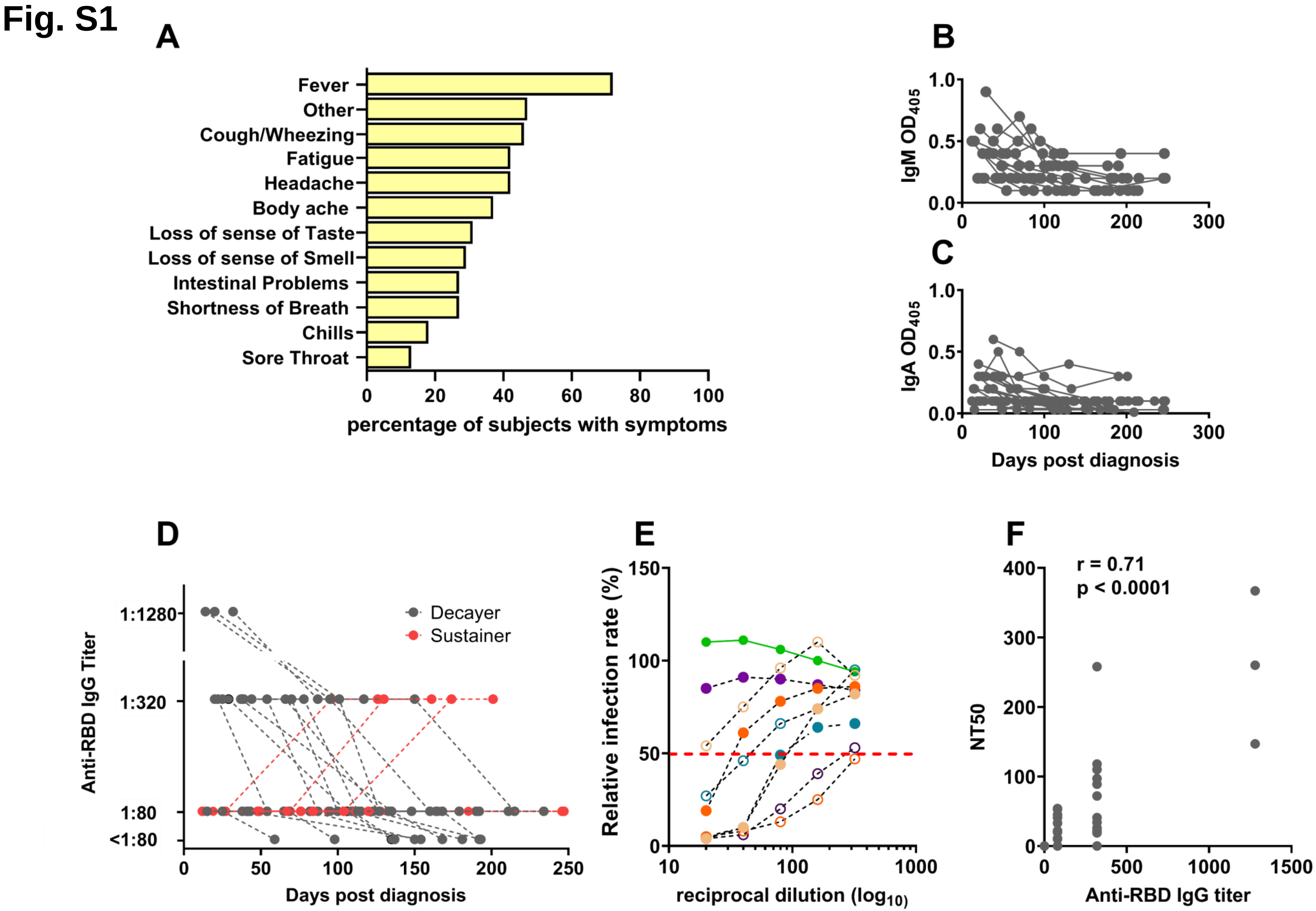
Fig. S1
A
Fever
Other
Cough/Wheezing
Fatigue
Headache
Body ache
Loss of sense of Taste
Loss of sense of Smell
Intestinal Problems
Shortness of Breath
Chills
Sore Throat
0
20
40
60
80
100
percentage of subjects with symptoms
B
IgM OD405
1.0
0.5
0.0
0
100
200
300
C
IgA OD405
1.0
0.5
0.0
0
100
200
300
Days post diagnosis
D
Anti-RBD IgG Titer
1:1280
1:320
1:80
<1:80
Decayer
Sustainer
0
50
100
150
200
250
Days post diagnosis
E
Relative infection rate (%)
150
100
50
0
10
100
1000
reciprocal dilution (log10)
F
NT50
r = 0.71
p < 0.0001
400
300
200
100
0
0
500
1000
1500
Anti-RBD IgG titer

**Supplementary Fig. 1. (A) Frequency of symptoms related to SARS-CoV-2 infection in study subjects (n = 83).** Each row represents a (type of) clinical symptom(s) and shows the percentage of study subjects who self-reported that symptom(s). **(B,C**) **Anti-RBD levels of IgM and IgA antibodies.** Line graphs showing IgM and IgA antibody levels in the same subjects as in main **Fig 2B**. (**D**) **Titers of circulating anti-RBD IgG.** Trajectories of RBD-specific IgG antibody titers corresponding to the $OD_{405}$ data in main **Fig. 1A.** Study subjects and definition of sustainers and decayers as per **Fig. 1B**. (**E**) **Method for neutralizing titer 50 (NT50) determination.** Representative neutralization curves demonstrating the determination of 50% neutralization (red dotted line) for the calculation of NT50 values presented in main **Fig. 1C** and main **Fig. 2C,D**. Relative infection rates were obtained by dividing the fluorescence readings (derived from fluorescently tagged virus) of the sample-treated wells by the un-treated control wells (infection rates above 100% result from small biological variations in untreated wells, and indicate no neutralization). The light green line corresponds to an uninfected (negative control) sample exhibiting no neutralizing activity. (**F**) **Correlation between anti-RBD titer and NT50.** Correlation analysis between anti-RBD IgG endpoint titers (x-axis) and neutralizing titers (y-axis) (n = 44). The Spearman's correlation coefficient r and corresponding p value were calculated.

Fig. S2

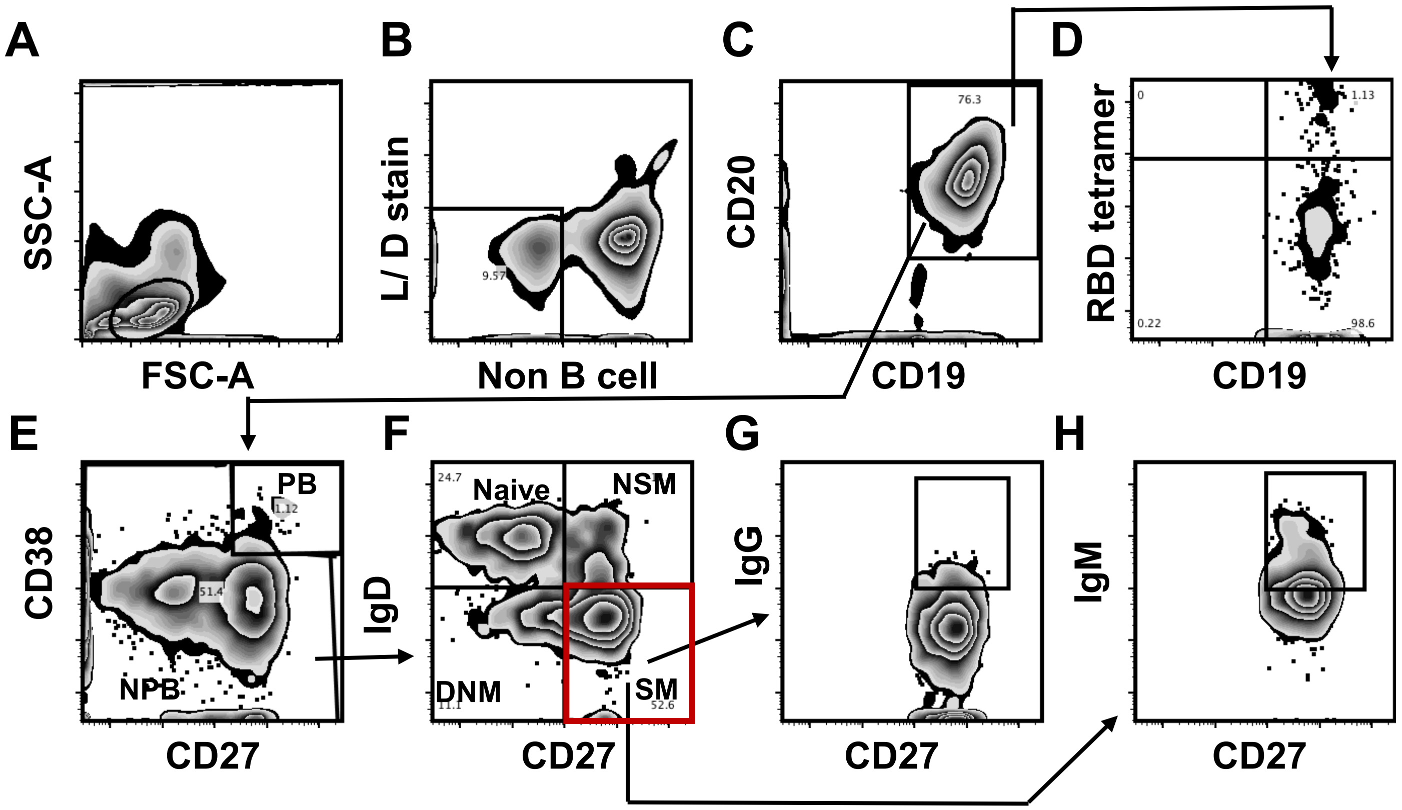

A
SSC-A
FSC-A
B
L/ D stain
9.57
Non B cell
C
CD20
76.3
CD19
D
RBD tetramer
0
1.13
0.22
98.6
CD19
E
CD38
PB
1.12
51.4
NPB
CD27
F
IgD
24.7
Naive
NSM
DNM
11.1
SM
52.6
CD27
G
IgG
CD27
H
IgM
CD27

**Supplementary Fig. 2. Flow cytometry gating strategy for B cell subsets.** (**A**) Physical parameters. (**B**) Exclusion of dead cells and non-B cells ($CD14^+$, $CD3^+$, $CD4^+$, $CD56^+$). (**C**) $CD19^+CD20^+$ cells (B cells) were further gated to distinguish (**D**) RBD-specific B cells utilizing fluorescently labeled RBD tetramers. (**E**) Plasmablast ($CD27^+CD38^{hi}$) and non plasmablasts. (**F**) Non-plasmablasts were further gated on CD27 and IgD for memory phenotyping: Naïve B cells ($CD27^-IgD^+$), Non switched memory B cells (NSM; $CD27^+IgD^+$), Switched memory B cells (SM; $CD27^+IgD^-$), and Double negative memory B cell (DNM; $CD27^-IgD^-$). Switched memory (SM) cell were further gated to determine the frequencies of (**G**) $IgG^+$ switched memory B cell ($IgG^+SM$) and (**H**) and $IgM^+$ switched memory B cell ($IgM^+SM$).

### Table S1. Characteristics of study participants

| Characteristics | n=83 |
|---|---|
| Age (years), median (IQR*) | 45 (31-56) |
| Gender, n (%) | |
| Male | 38 (46) |
| Female | 45 (54) |
| Race, n (%) | |
| African American or Black | 6 (7) |
| Alaskan Native of American Indian | 0 (0) |
| Asian or Pacific Islander | 13 (16) |
| White | 63 (76) |
| Other | 1 (1) |
| Ethnicity, n (%) | |
| Hispanic or Latino | 16 (19) |
| Non-Hispanic | 67 (81) |
| SARS-CoV-2 PCR Positivity, n (%) | |
| Positive | 81 (98) |
| Negative | 0 (0) |
| Not performed** | 2 (2) |
| Peak Disease Severity, n (%) [Female (F), Male (M)] | |
| Asymptomatic | 1 (1) [0F, 1M] |
| Mild (Non-hospitalized; 1-4 symptoms) | 55 (66) [30F, 25M] |
| Moderate (Non-hospitalized; 5 or more symptoms) | 22 (27) [16F, 6M] |
| Severe (Hospitalized) | 5 (6) [2F, 3M] |
| Days After Symptom Onset at Initial Collection (n=82***), median (IQR*) | 34 (26-42) |

*Interquartile range.
**Subjects who did not receive the PCR test were diagnosed by exposure history (PCR-positive household members), COVID-19 symptoms, and positive chest x-ray findings.
***The asymptomatic subject was not included.

### Table S2. Characteristics of study subjects followed longitudinally

| Characteristics | n=23 |
|---|---|
| Age (years), median (IQR*) | 39 (31-56) |
| Gender, n (%) | |
| Male | 9 (39) |
| Female | 14 (61) |
| Race, n (%) | |
| African American or Black | 2 (9) |
| Alaskan Native of American Indian | 0 (0) |
| Asian or Pacific Islander | 3 (13) |
| White | 18 (78) |
| Ethnicity, n (%) | |
| Hispanic or Latino | 6 (26) |
| Non-Hispanic | 17 (74) |
| SARS-CoV-2 PCR Positivity, n (%) | |
| Positive | 21 (91) |
| Negative | 0 (0) |
| Not performed** | 2 (9) |
| Peak Disease Severity, n (%) [Female (F), Male (M)] | |
| Asymptomatic | 1 (4) [0F, 1M] |
| Mild (Non-hospitalized; 1-4 symptoms) | 15 (65) [6F, 9M] |
| Moderate (Non-hospitalized; 5 or more symptoms) | 7 (31) [5F, 2M] |
| Severe (Hospitalized) | 0 (0) [0F, 0M] |
| Days After Symptom Onset at Initial Collection (n=22***), median (IQR*) | 38.5 (26-42) |

*Interquartile range.
**Subjects who did not receive the PCR test were diagnosed by exposure history (PCR-positive household members), COVID-19 symptoms, and positive chest x-ray findings.
***The asymptomatic subject was not included.

### Table S3. Comparison of cell subsets in non-infected and infected subjects

| Cell subset | Phenotype | A, Non-infected | B, Infected <2m post | C, Infected >5m post | p value | |
|---|---|---|---|---|---|---|
| | | | | | A vs. B [i] | B vs. C [ii] |
| Total B Cell | $CD19^+$ $CD20^+$ | 9.6 (7.0-12.0) | 6.0 (4.0-10.7) | 6.4 (4.8-8.1) | 0.08 | 0.36 |
| PB | $CD38^{hi}$ $CD27^+$ | 0.4 (0.2 - 0.7) | 1.1 (0.4-1.6) | 0.4 (0.2-0.9) | 0.01 | 0.04 |
| Naïve | $CD27^-$ $IgD^+$ | 55 (42.2-59.7) | 34.9 (25.9-34.8) | 30.2 (18.1-35.8) | 0.0004 | 0.04 |
| NSM | $CD27^+$ $IgD^+$ | 11 (6.9-16.2) | 12.1 (7.0-18.3) | 9.7 (2.8-14.6) | 0.27 | 0.04 |
| SM | $CD27^+$ $IgD^-$ | 23.4 (21.3-29.3) | 36.2 (26.6-51.6) | 35 (28.2-43.5) | 0.006 | 0.25 |
| SM $IgM^+$ | $CD27^+$ $IgD^-$ $IgM^+$ | 14.9 (9.2-22.3) | 32.7 (22.8-36.6) | 10.1 (7.7-17.3) | 0.001 | <0.0001 |
| SM $IgG^+$ | $CD27^+$ $IgD^-$ $IgG^+$ | 6.7 (2.0-8.0) | 4.3 (3.5-5.9) | 7.7 (2.7-14.9) | 0.21 | 0.04 |
| DNM | $CD27^-$ $IgD^-$ | 7.9 (5.0-12.5) | 10.4 (7.5-16.4) | 21.3 (12.2-34.6) | 0.08 | 0.003 |
| $RBD^+$ B Cell | $CD19^+$ $CD20^+$ $RBD^+$ | ND | 1.13 (0.88-1.36) | 1.03 (0.8-1.62) | NA | 0.08 |
| $RBD^+$ Naïve | $RBD^+CD27^-$ $IgD^+$ | ND | 50.9 (40.6-68.7) | 50 (43.3-62.0) | NA | 0.30 |
| $RBD^+$ NSM | $RBD^+$ $CD27^+$ $IgD^+$ | ND | 5.4 (3.3-14.3) | 8.6 (5.3-14.2) | NA | 0.4 |
| $RBD^+$ SM | $RBD^+$ $CD27^+$ $IgD^-$ | ND | 27.1(14.1-39.3) | 21.4 (13.2-35.6) | NA | 0.42 |
| $RBD^+$ DNM | $RBD^+$ $CD27^-$ $IgD^-$ | ND | 7.4 (4.4-11.1) | 11.8 (6.5-18.2) | NA | 0.01 |

Data are presented as median and interquartile range (IQR); PB - Plasmablast; NSM - non switched memory; SM - switched memory; DNM - double-negative memory

[i]Comparison between non-infected and infected (<2 months) using Mann-Whitney U test

[ii]Comparison between infected (<2 months) and infected (>5 months) using Wilcoxon matched pairs test

ND, Non detected

NA, Not applicable

### Table S4. Comparison of cell subsets in pre- and post-vaccinations of non-infected and infected subjects

| Cell subset | Phenotype | A, Infected, pre-vac | B, Infected, post-vac | C, Non-infected, pre-vac | D, Non-infected, post-vac | p value | |
|---|---|---|---|---|---|---|---|
| | | | | | | A vs. B [i] | C vs. D [ii] |
| B Cell | $CD19^+$ $CD20^+$ | 5.1 (3.3-7.2) | 3.5 (2.2 -4.7) | 12.3 (11.5-16.7) | 4.3 (3.2-4.8) | 0.03 | 0.004 |
| $RBD^+$ B Cell | $CD19^+$ $CD20^+$ $RBD^+$ | 0.5 (0.3-0.8) | 1.1 (0.9-1.6) | ND | 0.8 (0.5-0.9) | 0.003 | 0.004 |
| PB | $CD38^{hi}$ $CD27^+$ | 0.6 (0.1-1.1) | 0.4 (0-1.2) | 0.4 (0.06-0.9) | 0.2 (0.2-0.6) | 0.31 | 0.34 |
| Naïve | $CD27^-$ $IgD^+$ | 25.8 (12.5-37.1) | 63.3 (5.4-71.7) | 57.7 (49.1-63.6) | 66.5 (64.1-73.9) | 0.002 | 0.01 |
| NSM | $CD27^+$ $IgD^+$ | 5.7 (0.4-10.6) | 3.5 (2.7-9.3) | 9.7 (9.0-11.4) | 5.4 (3.7-8.9) | 0.42 | 0.01 |
| SM | $CD27^+$ $IgD^-$ | 41.7 (25.1-49.7) | 14.9 (12.5-17.7) | 23.4 (20.2-30.6) | 14.9 (12.5-17.7) | 0.002 | 0.01 |
| DNM | $CD27^-$ $IgD^-$ | 19.2 (13.3-49.8) | 14.7 (9.8-17.8) | 9.9 (2.9-10.6) | 9.8 (6.1-12.5) | 0.007 | 0.27 |

Data are presented as median and interquartile range (IQR); PB - Plasmablast; NSM - non switched memory; SM - switched memory; DNM - double negative memory; vac - vaccination

Comparisons of infected pre- vs post-vaccination[i] and non-infected pre- vs post-vaccination[ii] using Wilcoxon matched pairs test
